# Associations between vaping and Covid-19: cross-sectional findings from the HEBECO study

**DOI:** 10.1101/2020.12.01.20241760

**Authors:** Dimitra Kale, Aleksandra Herbec, Olga Perski, Sarah E Jackson, Jamie Brown, Lion Shahab

**Affiliations:** Department of Behavioural Science and Health, University College London, UK; Department of Clinical, Educational, and Health Psychology, University College London, UK

**Keywords:** vaping, e-cigarette, Covid-19, UK

## Abstract

**Aims:** To explore i) associations between vaping and self-reported diagnosed/suspected Covid-19; ii) changes in vaping since Covid-19 and factors associated with these changes; iii) whether Covid-19 motivated current or recent ex-vapers to quit.

**Methods:** Cross-sectional online survey of 2791 UK adults recruited 30/04/2020–14/06/2020. Participants self-reported data on sociodemographic characteristics, diagnosed/suspected Covid-19, vaping status, changes in vaping and motivation to quit vaping since Covid-19.

**Results:** There were no differences in diagnosed/suspected Covid-19 between never, current and ex-vapers. Bayes factors indicated there was sufficient evidence to rule out small negative (protective) associations between vaping status and diagnosed/suspected Covid-19. Among current vapers (n=397), 9.7% (95% CI 6.8-12.6%) reported vaping less than usual since Covid-19, 42.0% (37.2-46.9%) reported vaping more, and 48.3% (43.4-53.2%) reported no change. In adjusted analyses, vaping less was associated with being female (aOR=3.40, 95% CI 1.73-6.71), not living with children (aOR=4.93, 1.15-21.08) and concurrent smoking (aOR=8.77, 3.04-25.64), while vaping more was associated with being younger (aOR=5.26, 1.37-20.0), living alone (aOR=2.08, 1.14-3.85), and diagnosed/suspected Covid-19 (aOR=4.72, 2.60-8.62). Of current vapers, 32.2% (95% CI 27.5-36.8%) were motivated to quit vaping since Covid-19, partly motivated by Covid-19, and 17.4%, (9.7-26.3%) of recent ex-vapers quit vaping due to Covid-19.

**Conclusions:** Among UK adults, self-reported diagnosed/suspected Covid-19 was not associated with vaping status. Half of current vapers changed their vaping consumption since Covid-19, with the majority reporting an increase, and a minority was motivated to quit due to Covid-19.

**Registration:** The analysis plan was pre-registered, and it is available at https://osf.io/6j8z3/

**Highlights:**

- No difference found in diagnosed/suspected Covid-19 between never, current and ex-vapers
- Half of current vapers changed their vaping consumption since Covid-19
- Motivation to quit vaping was partly related to Covid-19

## 1. Introduction

The World Health Organization (WHO) declared the Covid-19 outbreak a global pandemic in March 2020. Covid-19 is an infectious disease that attacks the human respiratory system and can cause mild to severe illness and death (WHO, 2020). Using e-cigarettes, or vaping, is not harmless to lung function, but there has been little research on its impact on Covid-19. Tobacco smoking is more harmful to lungs than vaping (Denholm et al., 2010; Feldman & Anderson, 2013) but some research suggests that smokers may be at reduced risk of Covid-19 infection. While there remains uncertainty regarding the associated risk of Covid-19 hospitalisation, disease severity and mortality among smokers (Simons et al., 2020), some researchers have argued that the unexpected associations observed between smoking and Covid-19 outcomes may in part reflect either a protective effect of nicotine on Covid-19 initial infection, via interaction of nicotine with the ACE-2 receptor (Farsalinos et al., 2020), or on more severe disease progression after the infection, via the impact of nicotine on the anti-inflammatory cytokine pathway (Farsalinos et al., 2020; Kloc et al., 2020). In the absence of clear evidence, public health messaging has focused on the general benefits of quitting tobacco smoking (WHO, 2020; Public Health England, 2020). Some public health organisations have also urged people to stop vaping, claiming that vaping may increase susceptibility to Covid-19, despite little evidence to support this (Rizzo, 2020).

Irrespective of discussions surrounding potential protective effects of nicotine and whether there are substantial concerns about inhaling and exhaling vapour during a respiratory virus pandemic, it seems likely that the Covid-19 pandemic will have affected rates of vaping. On the one hand, an increase in attempts to stop smoking during the Covid-19 pandemic has been documented (Jackson et al, 2020), which could have increased uptake of vaping as a cessation aid. On the other hand, vape shops were shut during the UK lockdown, which might have reduced vaping. Data thus far are contradictory: some UK studies have reported reductions in vaping (Niedzwiedz et al., 2020), and increasing attempts to quit vaping due to concerns about Covid-19 (Tattan-Birch et al., 2020). Yet, another study in Italy, where vape shops were open during the lockdown, reported no change in vaping (Caponnetto et. al, 2020).

Given that e-cigarettes deliver nicotine without many of the harmful toxicants and carcinogens found in cigarette smoke (Shahab et al., 2017), investigating the association between vaping and Covid-19 infection may help delineate some of the proposed mechanisms for any potential protective or harmful effects of nicotine on Covid-19 outcomes. Further, understanding the impact of Covid-19 on vaping rates can help identify targets for intervention during future periods of social distancing and lockdown measures.

### 1.1 Aims

Using cross-sectional data from a survey of UK adults, this study aimed to investigate differences in diagnosed/suspected Covid-19 between vapers, ex-vapers and never vapers as well as changes in vaping during the Covid-19 pandemic and factors associated with these changes.

Specifically, we aimed to address the following research questions (RQs):

**RQ1**) Among adults in the UK, is vaping associated with self-reported diagnosed/suspected Covid-19, after adjustment for sociodemographic characteristics, smoking status and health conditions?

**RQ2**) Among current vapers, what proportion report vaping more, about the same, or less than usual since Covid-19; which characteristics are associated with changes, if any; what are the self-reported reasons for changes, and what proportion of current vapers are motivated to quit vaping because of Covid-19?

**RQ3**) Among recent ex-vapers (who stopped vaping ≤1 year ago), what proportion were motivated to quit vaping because of Covid-19; what proportion are considering taking up vaping again since Covid-19 and what are their self-reported reasons for this?

## 2. Methods

### 2.1 Study design

Analysis of cross-sectional data from the baseline wave of an ongoing longitudinal online study of UK adults; the HEalth BEhaviour during the COVID-19 pandemic (HEBECO) study (https://osf.io/sbgru/). Recruitment was online and through a number of channels including paid and unpaid advertisements on social media (including vaping forums) and through relevant mailing lists. HEBECO study data are collected and managed using REDCap electronic data capture tools hosted at University College London (Harris et al., 2009; Harris et al., 2019).

### 2.2 Study sample

UK-based adults aged 18 and over who completed the baseline survey of the HEBECO study between 30/04/2020 and 14/06/2020 (covering the period of the first national Covid-19-related lockdown in the UK). The UK Coronavirus Action Plan was published on 03/03/2020, followed by government advice to practice social distancing on 16/03/2020 and behavioural restrictions enforceable by law (‘lockdown’) on 23/03/2020 (Gov.uk, 2020). On 15/06/2020, most behavioural restriction measures were eased until the autumn.

### 2.3 Measures

For the full wording of all measures see Supplementary Materials 1.

#### 2.3.1 Outcomes

##### Diagnosed or suspected Covid-19

Participants were told that the key symptoms for Covid-19 are high temperature/fever or a new, continuous cough (consistent with UK official messaging about symptomatology at the time). They were then asked which of the following statements best applies to them: (i)I definitely have Covid-19 (I tested positive), (ii)I think I have Covid-19, (iii)I definitely had Covid-19 (I tested positive), (iv)I think I had Covid-19, (v)I do not have or think I have had Covid-19, (vi)Don’t know, and (vii)Prefer not to say. Those who replied (i, iii) were coded as ‘diagnosed’ and those who replied (ii, iv) as ‘suspected’ Covid-19 cases. For RQ1 we created a binary variable with answers (i) to (iv) were combined into ‘diagnosed/suspected Covid-19’ vs all other.

**Changes in vaping (current vapers only)** assessed with the question: ‘Has Covid-19 impacted on how much you vape?’ with the response options (i)I vape much less, (ii)I vape somewhat less, (iii)No change, (iv)I vape somewhat more, (v)I vape a lot more. We coded responses as: decrease vaping (i, ii), increase vaping (iv, v) and no change(iii).

**Reasons for changes in vaping (current vapers)** included the options: (1)Feeling stressed, (2)anxious/depressed, (3)relaxed, (4)lonely, (5)Out of boredom, (6)Changes in how much e-liquid I can buy, (7)Influence of others, (8)Staying mostly at home where there are fewer/no restrictions, (9)Other.

**Motivation to quit vaping (current vapers only)** assessed with the question ‘Which of the following best describes you?’ (adapted from Kotz, Brown & West, 2013) with the options ‘(i)I REALLY want to stop using e-cigs and intend to in the next month, (ii)I REALLY want to stop using e-cigs and intend to in the next 3 months, (iii)I want to stop using e-cigs and hope to soon, (iv)I REALLY want to stop using e-cigs but I don’t know when I will, (v)I want to stop using e-cigs but haven’t thought about when, (vi)I think I should stop using e-cigs but don’t really want to, (vii)I don’t want to stop using e-cigs’. Those who selected (i-v) were considered motivated to quit vaping.

**Reasons for motivation to quit vaping** included: (1)Rules around social distancing/self-isolation due to Covid-19, (2)Children/parents moved back home due to Covid-19, (3) Money is tighter due to Covid-19, (4)Decided it was too expensive, (5)Health problems/concerns related to Covid-19, (6)Health problems/concerns unrelated to Covid-19, (7)Advice from a GP, (8)Government/Tv/radio/press advert, (9)Social campaign, (10)Being contacted by local NHS Stop Smoking Services, (11)Being faced with restrictions already before Covid-19, (12)I knew someone else who was stopping, (13)Seeing a health warning on a packet, (14)Something said by family/friends/children, (15)Improve fitness, (16)Other. Participants who selected at least one of reasons 1,2,3, or 5 were classified as motivated to quit vaping for Covid-19-related reasons.

**Reasons for stopping vaping (recent ex-vapers only) (RQ3)** included the same options as described above in relation to reasons for motivation to quit vaping. Participants who selected at least one of the Covid-19-related reasons were classified as stopping vaping due to Covid-19-related reasons.

**Consideration of taking up vaping again (recent ex-vapers only) (RQ3)** dichotomized to no vs all other.

**Reasons for considering taking up vaping again (RQ3)** included: (1)Feeling stressed, (2)anxious/depressed, (3)relaxed, (4)lonely, (5)Out of boredom, (6)Struggling with cravings, (7)Influence of others, (8)To control weight, (9)Missing vaping, (10)Other.

#### 2.3.2 Explanatory variables

**Vaping status** was assessed with the question ‘Which statement about vaping best describes you?’ (adapted from Fidler et al., 2011). Participants were classified as current vapers (daily and non-daily), never vapers, ex-vapers (stopped vaping) and recent ex-vapers (stopped vaping ≤1 year). The latter two groups were not mutually exclusive.

#### 2.3.3 Covariates

**Heaviness of vaping (current vapers only)** calculated based on the following two questions (Heatherton et al., 1989): (i)’How many times per hour do you use your e-cigarette, on the days that you vape?’ (≤1(0); 1-5 times(1); 6-10 times(2); nearly all the time(3)) and (ii)’Usually, how soon after waking up do you draw your first puff on your e-cigarette, on the days that you vape?’ (≤5 minutes(3); 6-30 minutes(2); 31-60 minutes(1); >60 minutes(0)). Responses were dichotomized into low (scores 0-4) vs high (scores 5-6). **Sociodemographic information** included age in years (18-24, 25-34, 35-44, 45-54, 55-64, ≥65), gender (female vs all other), education (post-16 qualification vs not), household income pre-Covid-19 (<£50,000, ≥£50,000, prefer not to say; the selected cut-off reflects the median split in the analytic sample), ethnicity (White vs other), occupation (employed vs not), living alone (vs not), and living with children (vs not). Sociodemographic variables were included as covariates in our analyses given evidence that severe Covid-19 outcomes are disproportionately high among older, male, lower socioeconomic status and ethnic minorities (Public Health England, 2020; Whittle & Diaz-Artiles, 2020). Presence of any **health condition** (yes/no). Health condition was used as a covariate across analyses as people with health condition may be more likely to experience severe symptoms of Covid-19 (Onder, Rezza & Brusaferro, 2020; Wu & McGoogan, 2020). **Perceived risk of Covid-19** assessed by ‘What risk does Covid-19 pose to your health?’ and dichotomised to major/significant risk vs all other.

**Smoking status**, smokers (including smoking cigarettes or using any other tobacco product, daily or non-daily) vs not.

### 2.4 Statistical analysis

Analyses were conducted on complete cases using SPSS v.26. To account for the non-random nature of the sample, all data were weighted to the proportions of sex, age, ethnicity, education, household income and country of living obtained from the Office for National Statistics’ Annual Population Survey (2018).

**RQ1**: Logistic regression was used to examine associations between vaping status (never vapers (referent), ex-vapers and current vapers) and diagnosed/suspected Covid-19. We constructed three models: i)unadjusted; ii)with adjustment for sociodemographic characteristics; and iii)with additional adjustment for smoking status and health conditions. Where there were non-significant associations between vaping status and diagnosed/suspected Covid-19, Bayes factors (BFs) were used to distinguish inconclusive data from evidence for large, medium or small effects. Large effects in either direction were defined as having an odds ratio OR=4 (positive association) or for lower estimates 1/4 (negative association), based on evidence that current smoking rates among patients hospitalised for Covid-19 in China are four times lower than would be expected from the smoking prevalence in the country (Farsalinos, Barbouni & Niaura, 2020). We defined a medium effect size as OR=2 (positive association) or 1/2 (negative association) and a small effect size as OR=1.5 (positive association) or 2/3 (negative association).

**RQ2:** Among current vapers, we report changes in vaping since Covid-19 (i.e. the proportion who report vaping more, less, or about the same). We used multinomial logistic regression to examine associations of vaping characteristics, sociodemographic characteristics, perceived Covid-19 risk, health conditions, smoking status, and diagnosed/suspected Covid-19 with (i) vaping less, and (ii) vaping more, vs no change (referent). We analysed unadjusted associations (bivariate models) and independent associations (multivariable model) between each variable and outcome. Among current vapers who reported changes in vaping since Covid-19, we also report reasons for changes in vaping and calculated the proportion (and 95% Confidence Interval (CI)) of current vapers who are motivated to quit and describe their reasons (Covid-19-related vs. non-Covid-19-related) for considering stopping vaping.

**RQ3:** In recent ex-vapers (those who had stopped vaping ≤1 year ago), we calculated the proportion (and 95% CI) who reported quitting vaping due to Covid-19-related reasons. We also calculated the proportion (and 95% CI) of recent ex-vapers who reported considering taking up vaping again and their reasons for considering taking up vaping.

#### 2.4.1 Departures from the pre-registered protocol

The pre-registered protocol for the data analysis of RQ2 specified that logistic regressions would be used to examine associations of sociodemographic and vaping characteristics, perceived Covid-19 risk, health conditions, smoking status, and diagnosed/suspected Covid-19 with (i)vaping less (vs not [referent]) and (ii)vaping more (vs not [referent]). After analysing the data (reported on OSF, for transparency), we observed that a large proportion of vapers did not change their vaping since Covid-19. As a result, in a logistic regression the observed differences were primarily driven by the group who had not changed their vaping. Thus, we decided to conduct and report in the present manuscript additional analyses involving a multinomial regression analysis where the dependent variable had three levels: an increase, decrease and no change (reference category) in vaping status. Logistic regressions are presented in Supplementary Materials 2 (Table 4). We also amended the age categories to account for the observation that the number of participants in the category ≥70 years was small.

## 3. Results

A total of 2994 (weighted 2792) participants met the inclusion criteria for the study (complete cases) and were included in the present analyses. All the results presented below are weighted (for unweighted results see Supplementary Materials 2). Sample characteristics are shown in Table 1. Of the analytic sample, three quarters were never smokers and the rest were current or ex-vapers. Overall, one third of participants were aged 55-64 years, half were female, the majority were of White ethnicity, one third had no post-16 educational qualifications and three quarters had an annual income of less than £50,000.

**Table 1.** Sample characteristics (weighted data)

|  | Whole sample | Never vapers | Ex-vapers | Current vapers | *p*-value<sup>1</sup> |
| --- | --- | --- | --- | --- | --- |
| N (weighted) | 2792 | 2128 (76.2%) | 209 (7.5%) | 455 (16.3%) |  |
| Age in years, % (n) |  |  |  |  |  |
| 18-24 | 12.1 (338) | 10.5 (223) | 16.3 (34) | 17.8 (81) | <0.001 |
| 25-34 | 10.6 (295) | 10.4 (221) | 13.9 (29) | 9.9 (45) |  |
| 35-44 | 13.1 (367) | 12.6 (268) | 15.8 (33) | 14.5 (66) |  |
| 45-54 | 18.8 (526) | 16.9 (360) | 21.4 (45) | 26.6 (121) |  |
| 55-64 | 27.7 (774) | 29.0 (617) | 18.7 (39) | 25.9 (118) |  |
| $\geq 65$ | 17.6 (492) | 20.6 (439) | 13.9 (29) | 5.3 (24) | |
| Female sex, % (n) | 52.0 (1453) | 57.9 (1229) | 39.0 (82) | 31.1 (142) | <0.001 |
| White ethnicity, % (n) | 89.5 (2491) | 89.4 (1896) | 86.7 (182) | 91.2 (413) | 0.206 |
| Employed, % (n) | 47.3 (1322) | 47.4 (1008) | 44.8 (94) | 48.2 (220) | 0.702 |
| No post-16 qualifications, % (n) | 32.7 (913) | 31.7 (673) | 36.7 (77) | 35.7 (163) | 0.107 |
| Living alone, % (n) | 19.1 (533) | 17.8 (379) | 24.3 (51) | 22.6 (103) | 0.009 |
| Living with children, % (n) | 16.8 (469) | 16.0 (341) | 20.0 (42) | 18.9 (86) | 0.148 |
| Income, % (n) |  |  |  |  |  |
| ≥£50,000 | 17.8 (496) | 19.2 (409) | 11.8 (25) | 13.6 (62) | <0.001 |
| <£50,000 | 73.5 (2052) | 71.4 (1517) | 82.5 (174) | 79.2 (361) |  |
| Prefer not to say | 8.7 (243) | 9.3 (198) | 5.7 (12) | 7.2 (33) |  |
| Current tobacco smoker, % (n) | 26.0 (725) | 17.6 (375) | 70.5 (148) | 44.3 (202) | <0.001 |
| Health problems, % (n) | 44.3 (1214) | 43.4 (910) | 41.7 (85) | 49.5 (219) | 0.045 |
| Perceived high risk of Covid-19, % (n) | 26.9 (739) | 27.6 (579) | 29.3 (60) | 22.5 (100) | 0.068 |

<sup>1</sup>p values are for comparisons across the 3 subgroups

## 3.1 RQ1: Diagnosed/suspected Covid-19

Of the total sample, 1% (95% CI 0.7-1.4%) reported having a diagnosis of Covid-19 and a further 21.5% (95%CI 19.9-23.0%) reported suspected Covid-19. Table 2 shows the prevalence of diagnosed/suspected Covid-19 by vaping status. Both unadjusted and adjusted analyses showed no significant differences between vaping status and diagnosed/suspected Covid-19. The calculation of Bayes Factors (BFs) with expected effect sizes set to indicate a positive association (harmful effect) of ex- and current vaping compared with never vaping with diagnosed/suspected Covid-19 indicated that there was insufficient evidence to rule out small, medium (Supplementary Table 1) and large associations (Table 2). However, in adjusted analyses, there was sufficient evidence to rule out large differences in diagnosed/suspected Covid-19 for current compared with never vapers (Table 2). The calculation of BFs with expected effect sizes set to indicate a negative association (protective effect) of ex- and current vaping compared with never vaping with diagnosed/suspected Covid-19 indicated that there was sufficient evidence to rule out small, medium and large associations (Supplementary Table 2).

**Table 2.**
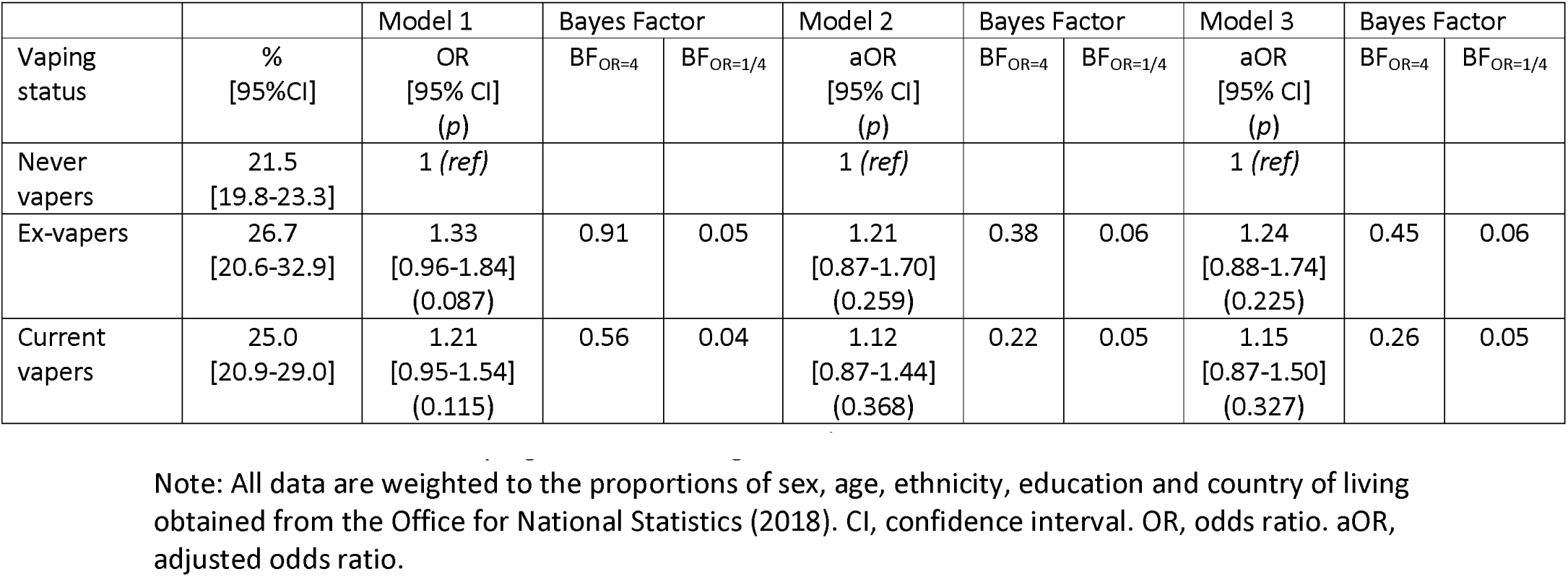
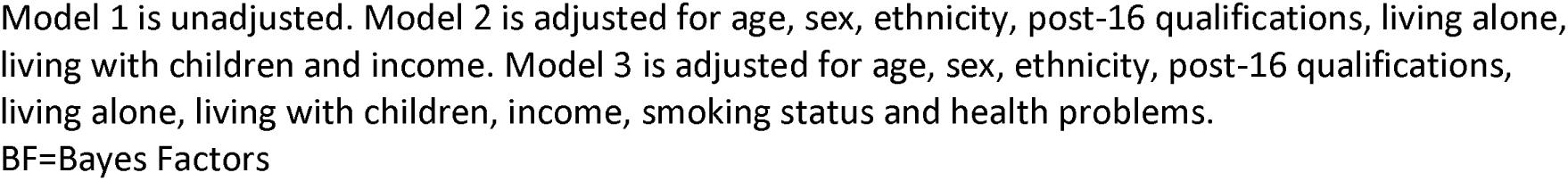
Associations of vaping status with diagnosed/suspected Covid-19

### 3.2 RQ2: Changes in vaping since Covid-19

Among current vapers, 38 (9.7%, 95%CI 6.8-12.6%) reported vaping less than usual since Covid-19, 167 (42.0%, 95%CI 37.2-46.9%) reported vaping more, and 192 (48.3%, 95%CI 43.4-53.2%) reported no change. Vaping less was independently associated with being female, not living with children and being a current tobacco smoker, while vaping more was independently associated with being younger, living alone, and diagnosed/suspected Covid-19 (Table 3).

**Table 3.** Correlates of vaping less and vaping more than usual since Covid-19 among current vapers n=397 (Multinomial logistic regression analyses)

|  | Vaping less n=38 |  |  |  | Vaping more n=167 |  |  |  |
| --- | --- | --- | --- | --- | --- | --- | --- | --- |
|  | Bivariate<br>Whole sample |  | Multivariable<br>Whole sample |  | Bivariate<br>Whole sample |  | Multivariable<br>Whole sample |  |
|  | OR<br>[95% CI] | *p* | aOR<br>[95% CI] | *P* | OR<br>[95% CI] | *p* | aOR<br>[95% CI] | *P* |
| Age 18-24 | 1 (*ref*) |  | 1 (*ref*) |  | 1 (*ref*) |  | 1 (*ref*) |  |
| 25-34 | 0.28<br>[0.06-1.33] | 0.110 | 0.27<br>[0.04-1.95] | 0.193 | 0.55<br>[0.24-1.27] | 0.164 | 0.34<br>[0.12-0.98] | **0.045** |
| 35-44 | 1.11<br>[0.37-3.31] | 0.857 | 2.36<br>[0.39-14.35] | 0.353 | 0.57<br>[0.25-1.28] | 0.171 | 0.24<br>[0.07-0.74] | **0.014** |
| 45-54 | 0.29<br>[0.10-0.88] | **0.029** | 0.42<br>[0.08-2.25] | 0.314 | 0.37<br>[0.19-0.73] | **0.004** | 0.25<br>[0.09-0.65] | **0.005** |
| 55-64 | 0.26<br>[0.08-0.80] | **0.019** | 0.28<br>[0.06-1.38] | 0.122 | 0.34<br>[0.17-0.68] | **0.002** | 0.22<br>[0.09-0.58] | **0.002** |
| ≥65 | 0.36<br>[0.07-2.04] | 0.250 | 0.88<br>[0.11-6.894] | 0.905 | 0.29<br>[0.10-0.87] | **0.027** | 0.19<br>[0.05-0.73] | **0.016** |
| Sex: Other | 1 (*ref*) |  | 1 (*ref*) |  | 1 (*ref*) |  | 1 (*ref*) |  |
| Female | 2.98<br>[1.47-6.04] | **0.003** | 4.13<br>[1.66-10.28] | **0.002** | 0.74<br>[0.47-1.18] | 0.208 | 1.00<br>[0.58-1.74] | 0.990 |
| Ethnicity: Other | 1 (*ref*) |  | 1 (*ref*) |  | 1 (*ref*) |  | 1 (*ref*) |  |
| White | 1<br>[0.29-3.42] | 0.999 | 1.55<br>[0.31-7.75] | 0.596 | 1.10<br>[0.52-2.34] | 0.803 | 1.67<br>[0.62-4.48] | 0.306 |
| Post 16 qualifications: yes | 1 (*ref*) |  | 1 (*ref*) |  | 1 (*ref*) |  | 1 (*ref*) |  |
| No | 0.91<br>[0.44-1.87] | 0.792 | 0.68<br>[0.25-1.84] | 0.449 | 0.78<br>[0.50-1.20] | 0.258 | 0.79<br>[0.45-1.37] | 0.395 |
| Employed: yes | 1 (*ref*) |  | 1 (*ref*) |  | 1 (*ref*) |  | 1 (*ref*) |  |
| Other | 0.64<br>[0.32-1.29] | 0.216 | 0.54<br>[0.18-1.60] | 0.265 | 0.97<br>[0.64-1.47] | 0.890 | 0.61<br>[0.35-1.05] | 0.075 |
| Living alone: yes | 1 (*ref*) |  | 1 (*ref*) |  | 1 (*ref*) |  | 1 (*ref*) |  |
| No | 1.58<br>[0.58-4.35] | 0.373 | 1.26<br>[0.32-4.98] | 0.741 | 0.54<br>[0.33-0.87] | **0.012** | 0.48<br>[0.26-0.88] | **0.018** |
| Living with children: yes | 1 (*ref*) |  | 1 (*ref*) |  | 1 (*ref*) |  | 1 (*ref*) |  |
| No | 1.96<br>[0.66-5.84] | 0.227 | 4.93<br>[1.15-21.08] | **0.032** | 0.84<br>[0.50-1.40] | 0.499 | 0.29<br>[0.09-0.93] | 0.347 |
| Income: ≥£50,000 | 1 (*ref*) | | 1 (*ref*) | | 1 (*ref*) | | 1 (*ref*) | |
| <£50,000 | 0.65<br>[0.25-1.72] | 0.385 | 0.98<br>[0.28-3.41] | 0.971 | 0.77<br>[0.42-1.43] | 0.409 | 0.92<br>[0.46-1.85] | 0.812 |
| Prefer not to say | 1.05<br>[0.25-4.36] | 0.952 | 0.80<br>[0.12-5.38] | 0.816 | 0.59<br>[0.21-1.63] | 0.307 | 0.49<br>[0.13-1.83] | 0.288 |
| Smoking status: Non-smoker | 1 (*ref*) |  | 1 (*ref*) |  | 1 (*ref*) |  | 1 (*ref*) |  |
| Smoker | 10.42<br>[4.20-26.32] | **<0.001** | 8.77<br>[3.04-25.64] | **<0.001** | 1.26<br>[0.82-1.93] | 0.299 | 0.84<br>[0.49-1.44] | 0.524 |
| Health problems: No | 1 (*ref*) |  | 1 (*ref*) |  | 1 (*ref*) |  | 1 (*ref*) |  |
| Yes | 0.84<br>[0.42-1.68] | 0.615 | 0.74<br>[0.28-1.93] | 0.533 | 0.90<br>[0.60-1.37] | 0.636 | 0.91<br>[0.54-1.55] | 0.739 |
| Perceived high risk of Covid-19: no | 1 (*ref*) |  | 1 (*ref*) |  | 1 (*ref*) |  | 1 (*ref*) |  |
| Yes | 1.30<br>[0.60-2.85] | 0.504 | 1.56<br>[0.58-4.26] | 0.380 | 1.04<br>[0.63-1.69] | 0.889 | 1.47<br>[0.81-2.67] | 0.208 |
| Diagnosed/suspected Covid-19: no | 1 (*ref*) |  | 1 (*ref*) |  | 1 (*ref*) |  | 1 (*ref*) |  |
| Yes | 1.61<br>[0.61-4.26] | 0.338 | 3.19<br>[0.96-10.64] | 0.059 | 5.65<br>[3.25-9.80] | **<0.001** | 4.72<br>[2.60-8.62] | **<0.001** |
| Light vaper | 1 (*ref*) |  | 1 (*ref*) |  | 1 (*ref*) |  | 1 (*ref*) |  |
| Heavy vaper | 0.02<br>[0-4.38] | 0.057 | 0.03<br>[0-1.94] | 0.101 | 1.20<br>[0.73-1.97] | 0.461 | 1.19<br>[0.66-2.13] | 0.565 |

Reference category: no change
Note: All data are weighted to the proportions of sex, age, ethnicity, education and country of living obtained from the Office for National Statistics (2018). CI, confidence interval. OR, odds ratio. aOR, adjusted odds ratio.
Multivariable model: age, sex, ethnicity, post-16 qualifications, living alone, living with children, income, smoking status, health problems, perceived risk of Covid-19 and confirmed/suspected Covid-19.
Missing data is due to attrition from the survey.

**Table 4.** Self-reported reasons for changes in vaping since Covid-19 in current vapers who reported changes since Covid-19 (n=205)

| Reason for changes in vaping | Vaping more (n=167)<br>% [95% CI] | Vaping less (n=38)<br>% [95% CI] |
| --- | --- | --- |
| Out of boredom | 56.9 [49.3-64.4] | 18.8 [5.9-31.8] |
| Staying mostly at home where there are fewer or no restrictions | 50.3 [42.6-57.9] | 23.5 [9.5-37.6] |
| Feeling anxious | 23.1 [16.7-29.6] | 18.9 [5.9-31.8] |
| Feeling stressed | 23.2 [16.7-29.6] | 13.1 [19.2-24.2] |
| Other | 9.6 [5.1-14.1] | 35.4 [19.6-51.3] |
| Feeling lonely | 14.4 [8.9-19.7] | 6.4 [0-14.5] |
| Feeling relaxed | 9.2 [4.8-13.7] | 4.4 [0-11.1] |
| Influence of other people | 6.5 [2.7-10.2] | 4.8 [0-11.8] |
| Changes in how much e-liquid /how many e-cigarettes I can buy | 2.1 [0-4.4] | 9.1 [0-18.6] |

Note: All data are weighted to the proportions of sex, age, ethnicity, education and country of living obtained from the Office for National Statistics (2018). CI, confidence interval.

The most common reason for vaping more was ‘out of boredom’, followed by ‘staying mostly at home where there are fewer/no restrictions’ (Table 4). Among vapers who reported vaping less, the most commonly specified reason was ‘staying at home where there are fewer/no restrictions’. In our sample, there were 128 (32.2%, 95% CI 21.7-37.9%) current vapers who reported being motivated to quit vaping due to Covid-19, 37 (29.8%, 95% CI 20.7-33.4%) of whom were motivated to do so for Covid-19-related reasons.

### 3.3 RQ3: Reasons for quitting vaping in the last year and considerations of taking up vaping again since Covid-19

There were 86 recent ex-vapers who reported reasons for quitting vaping and of these, 15 (17.4%, 95%CI 9.7-26.3%) had quit vaping due to Covid-19-related reasons. There were also 35 (40.7%, 95% CI 30.1-51.2%) recent ex-vapers who were considering taking up vaping again since Covid-19, and their most common reasons were ‘struggling with cravings’ (35.7%, 95% CI 22.9-48.5%) and ‘feeling stressed’ (25.2%, 95% CI 13.6-36.8%) (Supplementary Table 3).

## 4. Discussion

In this sample of UK adults, current vaping was not associated with diagnosed/suspected Covid-19. There was sufficient evidence to rule out a negative (i.e. protective) association between vaping status and diagnosed/suspected Covid-19, but the evidence was insufficient to rule out a small or medium positive (i.e. harmful) association. Half of current vapers changed their vaping consumption, with most of those who changed reporting an increase in their vaping since Covid-19 and doing so out of boredom or because they were staying home where there are fewer restrictions. One-third of current vapers were motivated to quit vaping since Covid-19; however, most of them were motivated to do so for non-Covid-19-related reasons. Our results also indicate that 17.4% of recent ex-vapers had quit vaping because of Covid-19, while 40.7% of recent ex-vapers were considering taking up vaping again since Covid-19.

Similar to research conducted in a representative sample of adults in England (Tattan-Birch et al., 2020), our findings indicate that there is no difference in self-reported Covid-19 infection between never, current and ex-vapers. In contrast, research on young adults (13-24 years old) in the US suggests that vaping is associated with a diagnosis of Covid-19 (Gaiha et al., 2020). This study has linked ever but not current vaping to Covid-19, and it was conducted among teens who are more prone than adults to take other risks related to Covid-19. Our study suggests that large effects of vaping on the risk of acquiring Covid-19 can be excluded but remains agnostic as to smaller effects.

It has been theorised that there may be a potential protective effect of nicotine on Covid-19 infection as there are biologically plausible pathways through which nicotine may impact Covid-19 cell entry (Farsalinos et al., 2020; Kloc et al., 2020). Our findings do not support such a protective effect. However, the current study was not able to address whether nicotine may be protective against the development of more severe symptoms of Covid-19 among people who become infected.

Similar proportions of current vapers reported vaping more or the same since Covid-19, and only a minority reported vaping less. Reduced vaping consumption was associated with being a smoker. This finding may indicate that dual users (those who are both cigarette smokers and vapers) have decreased their vaping since Covid-19, possibly because they instead increased their cigarette smoking. This is consistent with observed increases in cigarette consumption in current smokers in England since the start of the Covid-19 pandemic (Jackson et al., 2020). Our data also suggest that some people have reduced their vaping because of staying at home where there are fewer restrictions, with more opportunities to smoke cigarettes. Being female was also associated with reductions in vaping. During the first national UK lockdown, women reported being busier than ever with an increase in home-schooling, childcare, and care for older people (Engender, 2020). As such, women had less leisure time, which may result in vaping reductions.

Of current vapers, 42% reported an increase in vaping since Covid-19. We found that younger vapers were more likely than older vapers to report vaping more. Similarly, research in other addictive behaviours has found that younger people were more likely to increase their cigarette smoking and alcohol consumption since Covid-19 (Jackson et al., 2020; Garnett et al., 2020). Research has shown that younger people have been most affected by behavioural restrictions with large declines in their mental health and well-being (Etheridge & Spantig, 2020). Younger people have also reported worries about the impact of lockdown on their studies, work and relationships (ONS, 2020) and during lockdown, loneliness levels have been the highest and life satisfaction levels the lowest among younger adults (Fancourt & Steptoe, 2020). Indeed, our data indicated that the main reasons for increase in vaping were out of boredom and because people were staying at home.

Our analyses also showed that vapers with diagnosed/suspected Covid-19 had higher odds of reporting vaping more than usual since Covid-19, which may suggest a link between vaping consumption and Covid-19 infection. However, it should be highlighted that our data were cross-sectional and we did not prospectively examine this relationship. Thus, our findings may also suggest that vapers who believe they have/had Covid-19 started vaping more because of stress or believing that nicotine is protective against Covid-19. It is also possible, especially given the small number of confirmed cases of Covid-19 in our sample, that participants may have misinterpreted their symptoms as many other respiratory infections share symptoms with Covid-19.

One third of current vapers reported being motivated to quit vaping and were partly motivated to do so because of Covid-19. However, for most of vapers, their main reason for wanting to quit vaping was unrelated to Covid-19. This is in agreement with research conducted in Italy, which suggests that vapers did not consider stopping vaping during the first Italian lockdown (Caponnetto et al., 2020). It has also been documented that among cigarette smokers, attempts to quit were triggered by non-Covid-19 related reasons (Tattan-Birch et al., 2020). It is possible that contradictory media reports on the possibility of nicotine being protective against Covid-19 have partly discouraged people to quit vaping and smoking, while influencing recent ex-vapers to consider taking up vaping again. In our sample, 40.7% of recent ex-vapers were considering taking up vaping again since Covid-19 and one of the most common self-reported reasons was because of feeling stressed.

This study has a number of strengths. Data were collected in real-time during the first phase of the Covid-19 pandemic, which reduces the risk of recall bias. The variety of measures collected is another advantage, permitting a detailed analysis of a broad range of potential confounders of the relationship between vaping and diagnosed/suspected Covid-19, and correlates of changes in vaping since Covid-19. However, the study also had several limitations. First, diagnosed/suspected Covid-19 was self-reported and not confirmed with a viral or antibody test. As many other respiratory infections share symptoms with Covid-19, some participants may have misinterpreted their symptoms. In addition, it is likely that many participants experienced asymptomatic infection (Oran & Topol, 2020) and therefore did not report being infected. Second, our measure of vaping less likely underestimated the proportion reducing their vaping as those who quit vaping altogether since Covid-19 were not included in the analyses. Third, for some analyses the sample size was small, resulting in wide confidence intervals meaning that we likely lacked sufficient power to detect differences (confirmed by BFs, indicating data insensitivity). Fourth, while we weighted the sample to be representative of the UK adult population, it was self-selected and not random which affects the generalisability of the results. Fifth, most of the questions used were adapted from previous research and not validated, while changes in vaping were self-reported. Finally, the data were cross-sectional, and we did not measure the prospective relationship between changes in vaping and potential predictor variables. Longitudinal data following changes in vaping over time as the pandemic continues would be useful in evaluating the extent to which any initial changes in vaping behaviour are maintained over time.

## 4.1 Conclusions

When assessed by self-report in a UK population sample, diagnosed/suspected Covid-19 was not associated with vaping status. Among current vapers, half did not change their vaping consumption since Covid-19, with about 40% reporting an increase in vaping and 10% reporting a decrease. Vaping less was associated with being female, not living with children and concurrent smoking, while vaping more was associated with being younger, living alone, and diagnosed/suspected Covid-19. Motivation to quit vaping was partly related to Covid-19. A small number of recent ex-vapers quit vaping due to Covid-19, while nearly half of recent ex-vapers were considering taking up vaping again. Unlike cigarette smoking, there appears not to be a strong signal for any protective effect of vaping on diagnosed/suspected Covid-19. In addition, Covid-19 may have contributed to reinforcing different behavioural patterns (as observed for cigarette smoking and alcohol) such that a proportion of people have stopped completely since Covid-19, with others vaping more.

### Governance and ethics

The study has been approved by UCL Research Ethics Committee at the UCL Division of Psychology and Language Sciences (PaLS) (CEHP/2020/579) as part of the larger programme ‘The optimisation and implementation of interventions to change behaviours related to health and the environment’. All participants provided fully informed consent. The study is GDPR compliant.

## Data Availability

Data are available upon request

## Authors’ contributions

LS, AH, OP, DK and JB conceived and designed the study. DK and LS analysed the data. DK wrote the first draft. LS, AH, OP, SJ and JB provided critical revisions. All authors read and approved the submitted manuscript.

## Funding

DK, AH, OP and SJ receive salary support from Cancer Research UK (C1417/A22962). Funders had no role in the design and conduct of the study; collection, management, analysis and interpretation of the data; preparation, review or approval of the manuscript; and decision to submit the manuscript for publication.

## Competing interests

JB has received unrestricted research funding from Pfizer, who manufacture smoking cessation medications. LS has received honoraria for talks, an unrestricted research grant and travel expenses to attend meetings and workshops from Pfizer and has acted as paid reviewer for grant awarding bodies and as a paid consultant for health care companies. All authors declare no financial links with tobacco companies or e-cigarette manufacturers or their representatives.

## Supplementary Materials 1: Full Description of measures

### Outcomes

#### Diagnosed or suspected Covid-19 (RQ1)

Participants were told that the key symptoms for Covid-19 are high temperature/fever or a new, continuous cough (consistent with official symptomatology at the time). They were then asked which of the following statements best applies to them: (i) I definitely have Covid-19 (I tested positive), (ii) I think I have Covid-19, (iii)I definitely had Covid-19 (I tested positive), (iv) I think I had Covid-19, (v) I do not have or think I have had Covid-19, (vi) Don’t know, and (vii) Prefer not to say. Those who replied (i, iii) were coded as ‘diagnosed’ and those who replied (ii, iv) as ‘suspected’ Covid-19 cases.

**Changes in vaping (current vapers only) (RQ2)** assessed with the question: ‘Has Covid-19 impacted on how much you vape?’ with the response options (i) I vape much less, (ii) I vape somewhat less, (iii), No change, (iv) I vape somewhat more, (v), I vape a lot more. We coded decrease vaping (i, ii), increase vaping (iv, v) and no change(iii).

**Reasons for changes in vaping (current vapers) (RQ2)** Assessed with the question ‘Why have you changed how much you vape?’ with the options: (1) Feeling stressed, (2) Feeling anxious or depressed, (3) Feeling relaxed, (4) Feeling lonely, (5) Out of boredom, (6) Changes in how much e-liquid-how many e-cigarettes I can buy, (7) Influence of other people, (8) Staying mostly at home where there are fewer or no restrictions, (9) Other’.

**Motivation to quit vaping (current vapers only) (RQ2)** assessed with the question ‘Which of the following best describes you?’ (adapted from the Motivation To Stop Scale (Kotz, Brown & West, 2013)) with the options ‘(i) I REALLY want to stop using e-cigs and intend to in the next month, (ii) I REALLY want to stop using e-cigs and intend to in the next 3 months, (iii) I want to stop using e-cigs and hope to soon, (iv) I REALLY want to stop using e-cigs but I don’t know when I will, (v) I want to stop using e-cigs but haven’t thought about when, (vi) I think I should stop using e-cigs but don’t really want to, (vii) I don’t want to stop using e-cigs’. Those who selected (i-v) were considered motivated to quit vaping.

**Reasons for motivation to quit vaping (RQ2)** assessed with the question ‘What are the main reasons for you to consider stopping vaping now? (select all that apply)’ with the options: (1) Rules around social distancing or self-isolation due to Covid-19, (2) Children or parents moved back home due to Covid-19, (3) Money is tighter due to Covid-19, (4) Decided it was too expensive in general, (5) Health problems or concerns related to Covid-19, (6) Health problems or concerns unrelated to Covid-19, (7) Advice from a GP\health professional, (8) Government TV\radio\press advert, (9) Social campaign, e.g. on Twitter\advertisement of stop smoking treatment, (10) Being contacted by my local NHS Stop Smoking Services, (11) Being faced with smoking restrictions already before Covid-19, (12) I knew someone else who was stopping, (13) Seeing a health warning on a cigarette packet, (14) Something said by family\friends\children, (15) Improve my fitness, (16) Other. Participants who selected at least one of reasons 1,2,3, or 5 were classified as motivated to quit vaping for Covid-19-related reasons.

**Reasons for stopping vaping (recent ex-vapers only) (RQ3)** assessed with the question ‘What were the main reasons for you stopping vaping? (select all that apply)’ with the same options as described above in relation to reasons for motivation to quit vaping. Participants who selected at least one of the Covid-19-related reasons were classified as stopping vaping due to Covid-19-related reasons.

**Consideration of taking up vaping again (recent ex-vapers only) (RQ3)** assessed with the question ‘Are you considering taking up vaping again?’ with the options ‘(i) Definitely not, (ii) Probably not, (iii) Not sure, (iv) Probably yes, (v) Definitely yes’, dichotomized no(i, ii) vs all other (iii, iv, v).

**Reasons for considering taking up vaping again (RQ3)** assessed with the question ‘What are the main reasons for considering to go back to vaping? (select all that apply)’ with the options: (1) Feeling stressed, (2) Feeling anxious or depressed, (3) Feeling relaxed, (4) Feeling lonely, (5) Out of boredom, (6) I struggle with cravings, (7) Influence of other people around me, (8) To control weight, (9) I miss vaping, (10) Other, (11) Don’t know.

### Explanatory variables

#### Vaping status

Vaping status was assessed with the question ‘Which statement about vaping (e-cigarette use) best describes you?’ (Smoking in England, 2020), with the options i) I vape or use e-cigarettes every day; ii) I vape or use e-cigarettes but not every day; iii) I stopped vaping or using e-cigarettes completely in the last year; iv) I stopped vaping or using e-cigarettes completely more than a year ago; v) I have never vaped or used e-cigarettes. Those who select i) or ii) will be classified as current vapers, those who select iii) as recent ex-vapers, those who select iii) and iv) as long-term ex-vapers and those who select v) as never vapers.

### Covariates

**Heaviness of vaping (current vapers only)** calculated based on the following questions (Heatherton et al., 1989) (i) ‘How many times per hour do you use your e-cigarette, on the days that you vape?’ (≤1(0); 1-5 times(1); 6-10 times(2); nearly all the time(3)) and (ii) ‘Usually, how soon after waking up do you draw your first puff on your e-cigarette, on the days that you vape?’ (≤5 minutes(3); 6-30 minutes(2); 31-60 minutes(1); >60 minutes(0)). Responses were dichotomized low (scores 0-4) versus high (scores 5-6).

### Sociodemographic

**Age** categorical (≤24, 25-34, 35-44, 45-54, 55-64, ≥65)

Gender (female vs male/other)

**Education** 2 levels: [=0/1/2 vs all other] (0, No formal qualification | 1, GCSE/School certificate/O-level/CSE | 2, Vocational qualifications (e.g. NVQ1+2) VS. 3, A-level/Higher school certificate or equivalent (e.g. NVQ3) | 4, Bachelor degree or equivalent (e.g.NVQ4) | 5, Masters/ PhD/PGCE or equivalent | 6, Other)

**Household income pre-Covid**-19 3 levels: (<50 000/ ≥50 000/ unknown=prefer not to say) Ethnicity (2 levels; any white ethnicity vs all other including prefer not to say)

**Occupation** 2 levels: (employed (1, 2) versus not (all other) (1, Employed (full or part-time) | 2, Self-employed (full or part-time) | 3, Student | 4, Furloughed during Covid-19 | 5, Laid off during Covid-19 | 6, Unemployed since before Covid-19 | 7, Retired | 8, Homemaker, full-time parent or carer | 9, Unable to work due to disability | 10, Other)

**Living alone** measured with question “How many persons other than yourself (including children) live with you now in the same flat or house?”; Coded as 1= live alone, 0 = live with others).

**Living with Children** measured with the question “Do you live with any of these persons below?” (select all that apply) with the answers ‘with my partner’, ‘husband/wife’, ‘boyfriend/girlfriend’, ‘with children 0-5 years old’, ‘with children 6-15 years old’, ‘with persons aged 16-69 (family or friends)’, ‘with persons aged 70+ (family or friends)’, ‘persons who you believe may be vulnerable to Covid-19 for any reason’, ‘persons who are in poor health’. Living with children aged <16 coded as 1.

**Health condition**: 2 levels (yes vs no/prefer not to say): assessed by taking into account affirmative answers (‘yes’) to the following question ‘Do you have a health condition?’ Health condition was used as a covariate across analyses as people with health condition may be more likely to experience severe symptoms of Covid-19 (Onder, Rezza & Brusaferro, 2020; Wu & McGoogan, 2020).

**Perceived risk of Covid-19**: assessed by ‘What risk does Covid-19 pose to your health?’ and dichotomised to major/significant risk vs all other (Moderate risk, Minor risk, No risk at all, Don’t know).

**Smoking status**: was assessed with the question ‘Which statement about tobacco use and cigarette smoking best describes you?’ (Fidler et al., 2011), with the options i) I smoke cigarettes (including hand-rolled) every day; ii) I smoke cigarettes (including hand-rolled), but not every day; iii) I do not smoke cigarettes at all, but I do smoke tobacco of some kind (e.g. pipe, cigar or shisha); iv) I have stopped smoking completely in the last year; v) I stopped smoking completely more than a year ago; and vi) I have never smoked any cigarettes. Those who select i), ii) or iii) are classified as current smokers, those who select iv), v) or vi) as non-smokers.

## Supplementary Materials 2

**Table 1.** Bayes Factors for non-significant positive associations between vaping status and diagnosed/suspected Covid-19

|  | BF<br>large | BF<br>medium | BF<br>small | BF<sub>adj</sub><br>large | BF<sub>adj</sub><br>medium | BF<sub>adj</sub><br>small | BF<sub>adj</sub><br>large | BF<sub>adj</sub><br>medium | BF<sub>adj</sub><br>small |
| --- | --- | --- | --- | --- | --- | --- | --- | --- | --- |
| Never vapers | - | - | - | - | - | - | - | - | - |
| Ex-vapers | 0.91 | 1.68 | 2.38 | 0.38 | 0.71 | 1.07 | 0.45 | 0.86 | 1.29 |
| Current vapers | 0.56 | 1.07 | 1.66 | 0.22 | 0.44 | 0.71 | 0.26 | 0.52 | 0.82 |

Large expected effect sizes were set as OR=4, medium expected effect size as OR=2, small expected effect sizes as OR=1.5. BF=Bayes Factors, BF<sub>adj</sub>=Bayes Factors adjusted

**Table 2.**
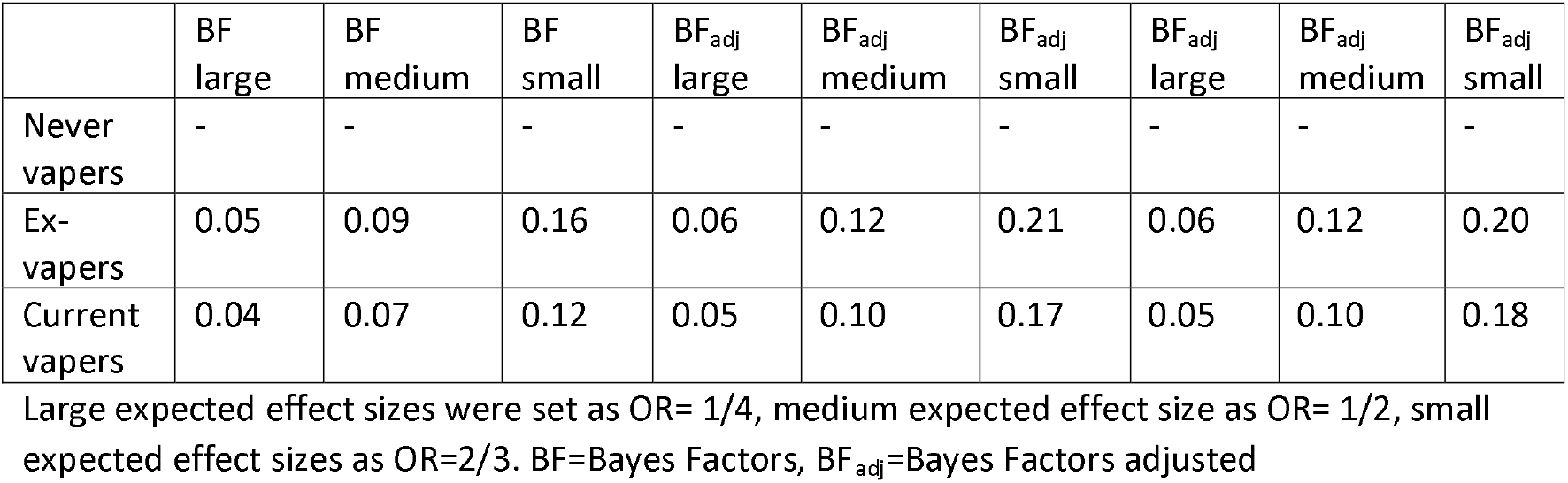
Bayes Factors for non-significant negative associations between vaping status and diagnosed/suspected Covid-19

**Table 3.** Self-reported reasons for considering taking up vaping again in recent ex-vapers n=35

| Reason for taking up vaping | % [95% CI] |
| --- | --- |
| I struggle with cravings | 35.7 [22.9-48.5] |
| Feeling stressed | 25.2 [13.6-36.8] |
| Other | 21.8 [10.8-32.9] |
| Feeling anxious | 14.9[5.4-24.4] |
| To control weight | 12.9 [4.0-21.9] |
| Out of boredom | 8.7 [1.2-16.2] |
| Feeling relaxed | 6.23 [0-12.7] |
| Miss vaping | 4.9 [0-10.7] |
| Influence of other people | 1.1 [0-3.9] |
| Feeling lonely | 0.43 [0-2.2] |

Note: All data are weighted to the proportions of sex, age, ethnicity, education and country of living obtained from the Office for National Statistics (2018). CI, confidence

**Table 4.**
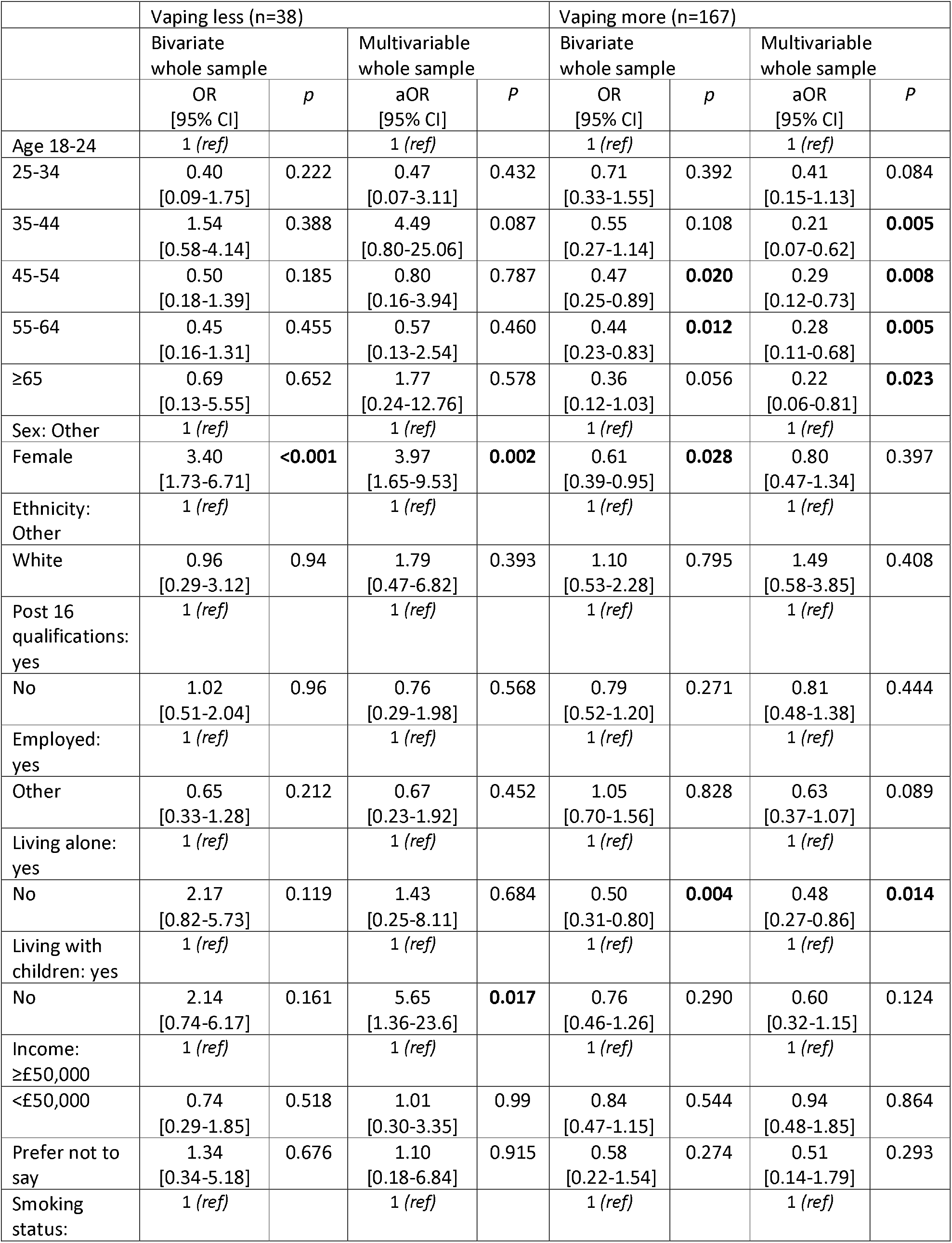
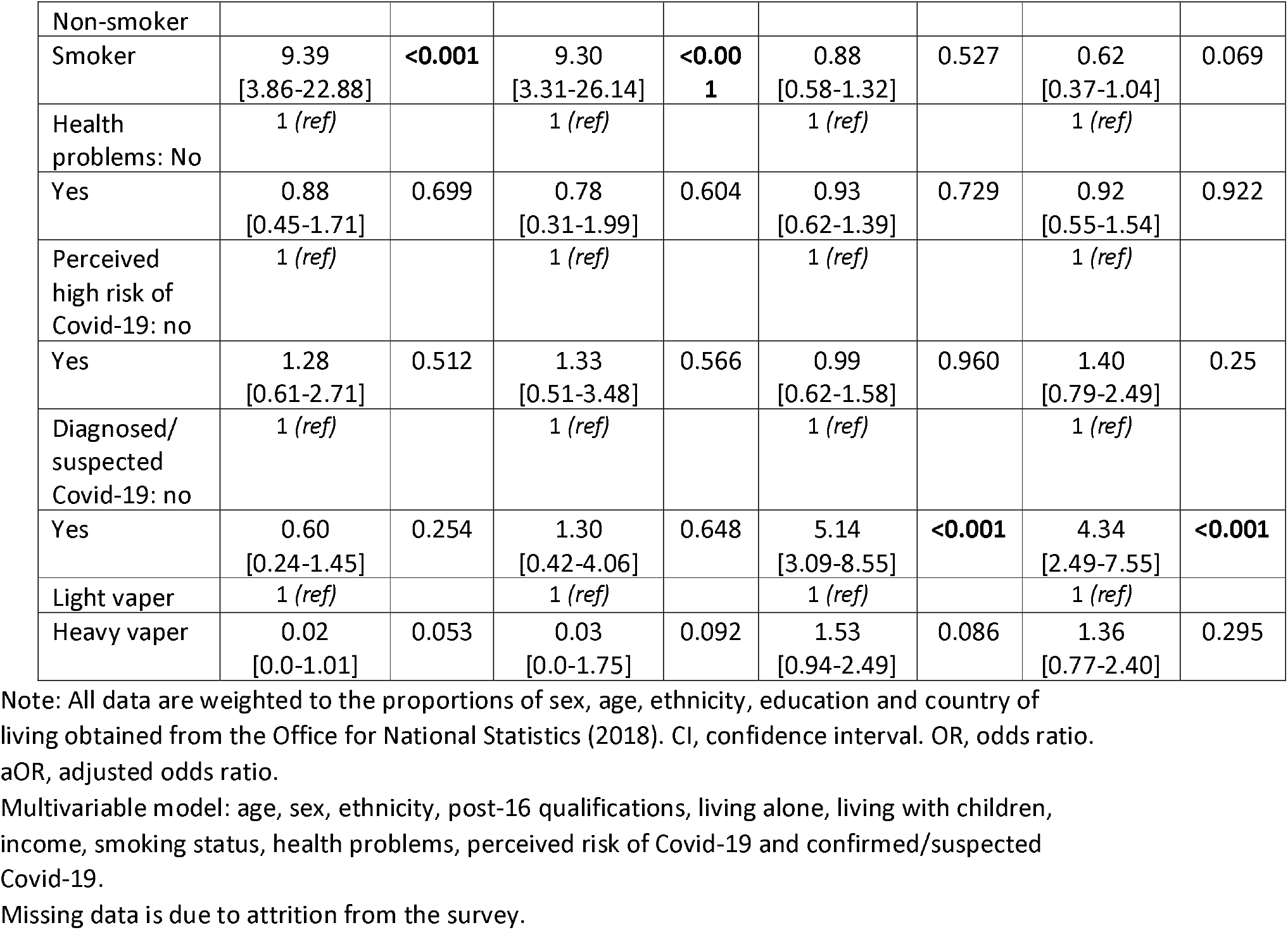
Correlates of vaping less and vaping more than usual since Covid-19 among current vapers (n=397) (Logistic regressions)

**Table 5.** Associations of vaping status with diagnosed/suspected Covid-19 unweighted analysis

| Vaping status | %<br>[95%CI] | Model 1<br>OR<br>[95% CI] | *p* | Model 2<br>aOR<br>[95% CI] | *p* | Model 3<br>aOR<br>[95% CI] | *p* |
| --- | --- | --- | --- | --- | --- | --- | --- |
| Never vapers | 20.7<br>[19.1-22.3] | 1 (ref) |  | 1 (ref) |  | 1 (ref) |  |
| Ex-vapers | 24.9<br>[18.4-31.4] | 1.27<br>[0.88-1.81] | 0.199 | 1.17<br>[0.81-1.69] | 0.396 | 1.12<br>[0.77-1.63] | 0.544 |
| Current vapers | 21.6<br>[17.1-26.1] | 1.05<br>[0.80-1.39] | 0.721 | 0.99<br>[0.74-1.32] | 0.931 | 0.94<br>[0.69-1.63] | 0.679 |

CI, confidence interval. OR, odds ratio. aOR, adjusted odds ratio.
Model 1 is unadjusted. Model 2 is adjusted for age, sex, ethnicity, post-16 qualifications, living alone, living with children and income. Model 3 is adjusted for age, sex, ethnicity, post-16 qualifications, living alone, living with children, income, smoking status and health problems.

**Table 6.** Correlates of vaping less and vaping more than usual since Covid-19 among current vapers (n=300) unweighted analysis; multinomial logistic regressions

|  | Vaping less n=40 |  |  |  | Vaping more n=123 |  |  |  |
| --- | --- | --- | --- | --- | --- | --- | --- | --- |
|  | Bivariate whole sample |  | Multivariable whole sample |  | Bivariate whole sample |  | Multivariable whole sample |  |
|  | OR<br>[95% CI] | *p* | aOR<br>[95% CI] | *p* | OR<br>[95% CI] | *p* | aOR<br>[95% CI] | *p* |
| Age 18-24 | 1 (*ref*) |  | 1 (*ref*) |  | 1 (*ref*) |  | 1 (*ref*) |  |
| 25-34 | 0.75<br>[0.19-3.03] | 0.687 | 0.33<br>[0.05-2.04] | 0.231 | 1.03<br>[0.36-2.94] | 0.958 | 0.41<br>[0.12-1.45] | 0.166 |
| 35-44 | 0.88<br>[0.27-2.89] | 0.836 | 0.80<br>[0.14-4.46] | 0.795 | 0.91<br>[0.36-2.31] | 0.839 | 0.30<br>[0.09-1.03] | 0.056 |
| 45-54 | 0.31<br>[0.10-1.00] | **0.049** | 0.26<br>[0.05-1.38] | 0.114 | 0.43<br>[0.18-1.00] | **0.050** | 0.15<br>[0.05-0.50] | **0.002** |
| 55-64 | 1.18<br>[0.05-0.63] | **0.007** | 0.14<br>[0.03-0.76] | **0.022** | 0.28<br>[0.12-0.66] | **0.004** | 0.12<br>[0.04-0.36] | **<0.001** |
| ≥65 | 0.45<br>[0.09-2.16] | 0.319 | 0.71<br>[0.10-4.92] | 0.728 | 0.23<br>[0.06-0.89] | **0.033** | 0.16<br>[0.03-0.79] | **0.024** |
| Sex: Other | 1 (*ref*) |  | 1 (*ref*) |  | 1 (*ref*) |  | 1 (*ref*) |  |
| Female | 2.27<br>[1.10-4.68] | **0.027** | 2.76<br>[1.12-6.80] | **0.027** | 0.99<br>[0.61-1.63] | 0.992 | 1.41<br>[0.79-2.51] | 0.241 |
| Ethnicity: Other | 1 (*ref*) |  | 1 (*ref*) |  | 1 (*ref*) |  | 1 (*ref*) |  |
| White | 2.91<br>[0.74-11.41] | 0.125 | 1.99<br>[0.38-10.60] | 0.418 | 1.34<br>[0.40-4.52] | 0.633 | 1.31<br>[0.30-5.75] | 0.721 |
| Post 16 qualifications: yes | 1 (*ref*) |  | 1 (*ref*) |  | 1 (*ref*) |  | 1 (*ref*) |  |
| No | 0.89<br>[0.37-2.14] | 0.797 | 1.13<br>[0.36-3.59] | 0.831 | 0.65<br>[0.35-1.23] | 0.186 | 0.71<br>[0.34-1.51] | 0.380 |
| Employed: yes | 1 (*ref*) |  | 1 (*ref*) |  | 1 (*ref*) |  | 1 (*ref*) |  |
| Other | 0.53<br>[0.26-1.07] | 0.078 | 0.6<br>[0.12-1.04] | 0.058 | 0.60<br>[0.36-0.97] | **0.039** | 0.47<br>[0.25-0.90] | **0.023** |
| Living alone: yes | 1 (*ref*) |  | 1 (*ref*) |  | 1 (*ref*) |  | 1 (*ref*) |  |
| No | 1.59<br>[0.61-4.13] | 0.343 | 1.43<br>[0.42-4.90] | 0.566 | 0.76<br>[0.43-1.35] | 0.355 | 0.62<br>[0.30-1.28] | 0.197 |
| Living with children: yes | 1 (*ref*) |  | 1 (*ref*) |  | 1 (*ref*) |  | 1 (*ref*) |  |
| No | 2.21<br>[0.72-6.74] | 0.164 | 4.79<br>[1.19-19.34] | **0.028** | 0.73<br>[0.41-1.31] | 0.289 | 0.74<br>[0.36-1.53] | 0.412 |
| Income: ≥£50,000 | 1 (*ref*) |  | 1 (*ref*) |  | 1 (*ref*) |  | 1 (*ref*) |  |
| <£50,000 | 0.54<br>[0.25-1.17] | 0.118 | 0.52<br>[0.19-1.40] | 0.195 | 0.62<br>[0.35-1.09] | 0.095 | 0.77<br>[0.40-1.52] | 0.454 |
| Prefer not to say | 0.43<br>[0.11-1.73] | 0.233 | 0.23<br>[0.04-1.33] | 0.101 | 0.35<br>[0.13-0.97] | **0.043** | 0.33<br>[0.09-1.16] | 0.083 |
| Smoking status: Non-smoker | 1 (*ref*) |  | 1 (*ref*) |  | 1 (*ref*) |  | 1 (*ref*) |  |
| Smoker | 8.77<br>[3.72-20.41] | **<0.001** | 5.85<br>[2.23-15.38] | **<0.001** | 1.09<br>[0.65-1.84] | 0.738 | 0.66<br>[0.35-1.25] | 0.202 |
| Health problems: No | 1 (*ref*) |  | 1 (*ref*) |  | 1 (*ref*) |  | 1 (*ref*) |  |
| Yes | 1.39<br>[0.69-2.82] | 0.357 | 1.60<br>[0.64-4.02] | 0.320 | 1.69<br>[0.63-1.69] | 0.894 | 1.22<br>[0.66-2.25] | 0.534 |
| Perceived<br>high risk of<br>Covid-19: No | 1 (*ref*) |  | 1 (*ref*) |  | 1 (*ref*) |  | 1 (*ref*) |  |
| Yes | 1.19<br>[0.52-2.70] | 0.680 | 1.56<br>[0.53-4.57] | 0.422 | 1.10<br>[0.62-1.97] | 0.747 | 1.77<br>[0.86-3.64] | 0.118 |
| Diagnosed/<br>suspected<br>Covid-19: No | 1 (*ref*) |  | 1 (*ref*) |  | 1 (*ref*) |  | 1 (*ref*) |  |
| Yes | 1.34<br>[0.48-3.68] | 0.576 | 1.91<br>[0.58-6.33] | 0.288 | 3.65<br>[1.92-6.94] | **<0.001** | 3.36<br>[1.66-6.80] | **0.001** |
| Light vaper | 1 (*ref*) |  | 1 (*ref*) |  | 1 (*ref*) |  | 1 (*ref*) |  |
| Heavy vaper | 0.09<br>[0.01-0.66] | **0.019** | 0.13<br>[0.02-1.11] | 0.062 | 1.16<br>[0.65-2.08] | 0.611 | 1.16<br>[0.60-2.26] | 0.656 |

Reference category: no change
CI, confidence interval. OR, odds ratio. aOR, adjusted odds ratio.
Multivariable model: age, sex, ethnicity, post-16 qualifications, living alone, living with children, income, smoking status, health problems, perceived risk of Covid-19 and confirmed/suspected Covid-19.

## Notes

### Author Declarations

The study has been approved by UCL Research Ethics Committee at the UCL Division of Psychology and Language Sciences (PaLS) (CEHP/2020/579) as part of the larger programme: The optimisation and implementation of interventions to change behaviours related to health and the environment. All participants provided fully informed consent. The study is GDPR compliant.

